# Development and validation of the Chinese Osteoporosis Screening Algorithm (COSA) in Identification of People with High Risk of Osteoporosis

**DOI:** 10.1101/2022.10.20.22281292

**Authors:** Ching-Lung Cheung, Gloria Hoi-Yee Li, Constance Mak, Kathryn CB Tan, Annie WC Kung

## Abstract

Osteoporosis is a prevalent but under-diagnosed disease partly due to the limited availability of dual-energy X-ray absorptiometry (DXA) machines in many countries. To enhance the public awareness and facilitate diagnosis of osteoporosis, we aimed to develop a new Chinese Osteoporosis Screening Algorithm (COSA) and compare its performance with the Osteoporosis Self-Assessment Tool for Asians (OSTA) to identify people at high risk of osteoporosis. A total of 4,747 postmenopausal women and men aged≥50 years from the Hong Kong Osteoporosis Study were randomly split into a development (N=2,373) and an internal validation cohort (N=2,374). An external validation cohort comprising 1,876 community-dwelling subjects was used to evaluate the positive predictive value (PPV). Among 11 predictors included, age, sex, weight, and history of fracture were significantly associated with osteoporosis after correction for multiple testing in the development cohort. Age- and sex-stratified models were developed separately for women aged<65 years, women aged≥65 years, and men due to the presence of significant sex and age interactions. The AUC of the COSA in the internal validation cohort was 0.761 (95% CI: 0.711-0.811), 0.822 (95% CI: 0.792-0.851), and 0.946 (95% CI: 0.908-0.984) for women aged<65 years, women aged≥65 years, and men, respectively. The COSA outperformed OSTA in identifying people with osteoporosis in all the three groups. In the external validation cohort, the PPV of COSA was 40.6%, 59.4%, and 19.4% for women aged<65 years, women aged≥65 years, and men, respectively. In conclusion, we have developed and validated a new osteoporosis screening algorithm, COSA, specific for Hong Kong Chinese.

## Introduction

Osteoporosis is a prevalent disease affecting hundred millions of people worldwide. We recently projected the hip fracture number in Asia and found that the number of hip fracture will be doubled by 2050.^1^ Thus, there is an urgent need to reduce the incidence of hip fracture, particularly in Asia.

Although osteoporosis is a public health issue, it is widely recognized as an underdiagnosed disease partly due to the limited availability of dual-energy X-ray absorptiometry (DXA), which is the gold standard of bone mineral density (BMD) measurement. For example, the minimum number of DXA machine required for adequate osteoporosis care was established to be 11 per million people.^2^ However, many parts of the world have insufficient DXA machines.^3^ In particular, people living in the rural areas have restricted access to DXA. Thus, to facilitate early diagnosis of osteoporosis, it is important to develop a simple tool that can help prioritizing people to have DXA scan.

Risk prediction tool is important in guiding clinical management. Previously, Osteoporosis Self-Assessment Tool for Asians (OSTA) was developed by a group of experts to identify Asian women who are at high-risk of osteoporosis, using just age and weight as the predictor variables.^4^ OSTA was subsequently validated in men.^5^ However, the the OSTA was developed based on women from different Asian countries and regions. It is now widely recognized that population-specific risk factors are important in developing risk prediction tools. Most importantly, the estimates of the risk factors used in the prediction model must be derived from the population that the model applies. In this study, we aimed to develop a new Chinese Osteoporosis Screening Algorithm (COSA) and compare its performance with the OSTA.

## Materials and methods

### Development and internal validation cohort

In this study, we used the data from the Hong Kong Osteoporosis Study (HKOS) for the development and internal validation. Details of the HKOS has been previous described.^6^ In this study, 6,120 post-menopausal women and men aged 50 years or above were included. After excluding participants with missing data on femoral neck BMD and/or other predictors (N=1,373), 4,747 participants were included in the final analysis. These participants were randomly split into a development (N=2,373) and an internal validation cohort (N=2,374).

### External validation cohort

We recruited and screened 2,012 community dwelling participants aged 50 years or above using the COSA questionnaire during 2019-2021 in Kwai Tsing District in Hong Kong. We further invited the high-risk participants identified using the optimal cutoff point of COSA, for DXA scanning.

### Variables included in the prediction model development

We selected the risk factors included in the FRAX (age, sex, height, weight, history of fracture, smoking, drinking, use of steroid, and rheumatoid arthritis) except parental history of fracture and secondary osteoporosis. Since parental history of fracture was only available in less than half of the participants, including this variable in the model would largely reduce the sample size. While secondary osteoporosis is a broad term, we included in the model two disease predictors (diabetes and stroke) that are closely related to fracture instead. Diabetes is a well-established risk factor of fracture,^7^ and our recent hip fracture prediction score^8^ showed that stroke was a significant predictor of hip fracture in Hong Kong Chinese.

### OSTA

In the validation cohort, we compared the performance of COSA with OSTA in identification of people with osteoporosis. OSTA index was calculated using the following formula: OSTA = 0.2 * [weight (kg) - age (years)]. OSTA<-1 indicated intermediate and high risk of osteoporosis.^4^

### Clinical outcomes

Osteoporosis was defined as BMD T-score ≤ −2.5 at the femoral neck. BMD was measured using DXA (Hologic QDR 4500 plus). Since BMD at the femoral neck is the most robust predictor of hip fracture, osteoporosis at the femoral neck is used as the primary outcome in the COSA development. The secondary outcome was osteoporosis at either spine or hip (lumbar spine, femoral neck, or total hip).

### Statistical analysis

The COSA was developed using logistic regression, with bias-corrected accelerated 95% Confidence Interval (CI) and p-value estimated using 1,000 bootstrap resamples. Variables showing significant association with osteoporosis after correction for multiple testing (0.05/11=0.0046) were selected to build the COSA model. We evaluated the model interaction with age and sex and found significant interaction, therefore we eventually developed the COSA by obtaining the estimates of the variables in three groups separately: (1) women aged<65 years; (2) women aged≥65 years; and (3) men. Men was not further divided by age because the limited sample size. The area under the receiver operating characteristic (ROC) curve (AUC) of the COSA and OSTA were determined. To compare the improvement in classification of osteoporosis of COSA with reference to OSTA, category-less net reclassification index (NRI) and integrated discrimination improvement (IDI) were evaluated using the R package “Hmisc”.

The Youden’s index was used to select the cutoff point of the COSA to identify people at high risk of osteoporosis. The final COSA equations were developed by scaling of the beta-coefficient of each variable and the incorporation of the Youden’s index, such that an individual with COSA>0 is at high-risk of osteoporosis. The sensitivity, specificity, positive predictive value (PPV), and negative predictive value (NPV) of the Youden’s index were calculated. Association with p-value <0.05 was considered statistically significant. All analyses were conducted using SPSS version 22.0 software (IBM, Inc., Chicago, IL) and R (R Foundation for Statistical Computing, Vienna, Austria; https://www.r-project.org/).

### Ethics

The study protocol was approved by the institutional review board of the University of Hong Kong and the HA Hong Kong West Cluster (reference: UW03-140 and UW 20-333).

### Role of the funding source

The funders were not involved in the study design, collection, analysis, and interpretation of data, nor did they have a role in the writing of the manuscript and decision to submit it for publication. All authors had full access to all the data in the study and accepted the responsibility to submit it for publication.

## Results

### Development of COSA

Table 1 shows the characteristics of the study participants included in the development and internal validation cohort. The characteristics were similar between the development and internal validation cohorts, except that the development cohort had a high prevalence of rheumatoid arthritis.

**Table 1.** Demographic characteristics of the HKOS study participants.

|  | Development cohort |  | Internal validation cohort |  | P-value |
| --- | --- | --- | --- | --- | --- |
|  | Mean or<br>N | SD or<br>% | Mean or<br>N | SD or<br>% |  |
| Age | 65.70 | 9.50 | 65.80 | 9.40 | 0.706 |
| Height (m) | 1.56 | 0.09 | 1.56 | 0.09 | 0.267 |
| Weight (kg) | 57.32 | 10.55 | 57.06 | 10.42 | 0.395 |
| Female | 1573 | 66.3 | 1595 | 67.2 | 0.511 |
| Ever smoking | 396 | 16.7 | 355 | 15.0 | 0.102 |
| Ever drinking | 308 | 13.0 | 270 | 11.4 | 0.092 |
| History of fracture | 579 | 24.4 | 594 | 25.0 | 0.62 |
| Diabetes | 308 | 13.0 | 276 | 11.6 | 0.156 |
| Stroke | 49 | 2.1 | 47 | 2.0 | 0.835 |
| Use of oral steroids | 37 | 1.6 | 24 | 1.0 | 0.096 |
| Rheumatoid arthritis | 16 | 0.7 | 5 | 0.2 | 0.023 |
| BMD at the femoral neck<br>(g/cm<sup>2</sup>) | 0.634 | 0.129 | 0.630 | 0.129 | 0.325 |
| BMD T-score at the femoral<br>neck | -1.47 | 1.15 | -1.50 | 1.14 | 0.302 |
| Osteoporosis at the femoral<br>neck | 451 | 19.0 | 457 | 19.3 | 0.83 |

Continuous variables are presented as mean and standard deviation (SD). Categorical variables are presented as number (N) and %.

Table 2 shows the result of the association of the risk factors with osteoporosis. In the multivariable logistic regression analysis, sex, age, weight, and history of fracture were significantly associated with osteoporosis after correction for multiple testing. Similar significant associations of these factors with osteoporosis were also observed in the internal validation cohort (Table 2). Therefore, these variables were used for the risk score development. In the risk score developed based on the beta-estimate of these variables, we found that there was a significant sex and age interaction with the risk score (P_interaction_<0.05), therefore we developed the risk score in three groups separately, namely women aged<65 years, women aged≥65 years, and men.

**Table 2.** Multivariable logistic regression in the development and internal validation cohorts.

| Variables used in the<br>model development | Development cohort |  |  |  | Internal validation cohort |  |  |  |
| --- | --- | --- | --- | --- | --- | --- | --- | --- |
|  | OR | 95% CI |  | P-value | OR | 95% CI |  | P-value |
|  |  | lower | Upper |  |  | lower | Upper |  |
| Age (years) | 1.128 | 1.106 | 1.154 | 0.001 | 1.116 | 1.097 | 1.139 | 0.001 |
| Height (m) | 0.195 | 0.012 | 3.144 | 0.234 |  |  |  |  |
| Weight (kg) | 0.901 | 0.880 | 0.919 | 0.001 | 0.879 | 0.857 | 0.896 | 0.001 |
| Female | 15.662 | 8.681 | 35.200 | 0.001 | 12.787 | 7.486 | 26.364 | 0.001 |
| Ever smoking | 1.448 | 0.791 | 2.488 | 0.195 |  |  |  |  |
| Ever drinking | 0.919 | 0.456 | 1.738 | 0.804 |  |  |  |  |
| History of fracture | 2.510 | 1.827 | 3.512 | 0.001 | 2.809 | 2.075 | 3.838 | 0.001 |
| Diabetes | 0.770 | 0.513 | 1.188 | 0.212 |  |  |  |  |
| Stroke | 0.987 | 0.283 | 3.292 | 0.986 |  |  |  |  |
| Use of oral steroids | 0.951 | 0.237 | 2.944 | 0.924 |  |  |  |  |
| Rheumatoid arthritis | 6.021 | 1.155 | 26.053 | 0.009 |  |  |  |  |

95% CI and p-value were computed based on 1,000 bootstrap samples.

After deriving the COSA score and identifying the Youden’s index in each group, the beta coefficient of each variable was scaled and Youden’s index was incorporated into the COSA equation, so that COSA score>0 indicates a high risk of osteoporosis. The final equation of the COSA in three groups are provided in Table 3. The AUC of the COSA in the development cohort was 0.810 (95% CI: 0.762-0.859), 0.807 (95% CI: 0.776-0.838), and 0.923 (95% CI: 0.860-0.985) for women aged<65 years, women aged≥65 years, and men, respectively (Supplementary Table 1 and Supplementary Figure 1). For osteoporosis at either spine or hip, the AUC of the COSA are provided in Supplementary Table 1.

**Table 3.** The original and scaled beta-coefficient for COSA

|  | Women aged<65 years |  | Women aged≥65 years |  | Men |  |
| --- | --- | --- | --- | --- | --- | --- |
|  | Beta | Scaled beta | Beta | Scaled beta | Beta | Scaled beta |
| Age (years) | 0.105 | 3 | 0.099 | 2 | 0.077 | 1 |
| Weight (kg) | -0.143 | -4 | -0.092 | -2 | -0.151 | -2 |
| History of fracture | 1.057 | 32 | 0.814 | 17 | 1.308 | 17 |
| Constant | -1.237 | 17<sup>#</sup> | -2.771 | -53<sup>#</sup> | -1.036 | 34<sup>#</sup> |
| Equation | 17 + (age *3) + (weight * -4)<br>+ (history of fracture * 32) |  | -53 + (age * 2) + (weight * -<br>2) + (history of fracture * 17) |  | 34 + (age* 1) + (weight * -2)<br>+ (history of fracture * 17) |  |

<sup>#</sup>The scaled beta of the constant included the Youden's index, such that the COSA score > 0 indicates a high risk of osteoporosis.

### Internal validation of COSA

In the internal validation cohort, we compared the performance of COSA with reference to OSTA in risk stratification of osteoporosis (Table 4). In all the three groups, the COSA had higher AUC (Figure 1) and accuracy than OSTA in the identification of subjects with high risk of osteoporosis at the femoral neck (Table 4). Using IDI and category-less NRI, COSA had a significant improvement in the reclassification of osteoporosis compared to OSTA in all the three groups (Table 5; all P<0.05). Similar significant improvement was observed in reclassification of osteoporosis at either spine or hip (data not shown).

**Table 4.** The risk stratification of COSA and OSTA in the internal validation cohort in (a) women aged<65 years, (b) women aged≥65 years, and (c) men.

|  | COSA |  | OSTA |  |
| --- | --- | --- | --- | --- |
|  | Value | 95% CI | Value | 95% CI |
| AUC | 0.761 | 0.711 to 0.811 | 0.736 | 0.685 to 0.787 |
| Sensitivity | 57.47% | 46.41% to 68.01% | 67.82% | 56.94% to 77.44% |
| Specificity | 78.16% | 75.05% to 81.05% | 63.55% | 60.02% to 66.98% |
| PPV | 23.15% | 19.38% to 27.40% | 17.56% | 15.20% to 20.20% |
| NPV | 94.14% | 92.61% to 95.36% | 94.52% | 92.68% to 95.92% |
| Accuracy (*) | 76.03% | 73.01% to 78.87% | 63.99% | 60.65% to 67.23% |

(b) Women aged≥65 years
|  | COSA |  | OSTA |  |
| --- | --- | --- | --- | --- |
|  | Value | 95% CI | Value | 95% CI |
| AUC | 0.822 | 0.792 to 0.851 | 0.814 | 0.783 to 0.844 |
| Sensitivity | 67.16% | 61.87% to 72.14% | 97.34% | 95.01% to 98.78% |
| Specificity | 80.00% | 75.80% to 83.77% | 18.05% | 14.45% to 22.12% |
| PPV | 73.46% | 69.23% to 77.31% | 49.47% | 48.26% to 50.69% |
| NPV | 74.72% | 71.58% to 77.62% | 89.16% | 80.69% to 94.18% |
| Accuracy (*) | 74.20% | 70.90% to 77.30% | 53.88% | 50.23% to 57.49% |

(c) Men
|  | COSA |  | OSTA |  |
| --- | --- | --- | --- | --- |
|  | Value | 95% CI | Value | 95% CI |
| AUC | 0.946 | 0.908 to 0.984 | 0.930 | 0.884 to 0.976 |
| Sensitivity | 90.62% | 74.98% to 98.02% | 96.88% | 83.78% to 99.92% |
| Specificity | 86.08% | 83.39% to 88.48% | 53.28% | 49.63% to 56.91% |
| PPV | 21.80% | 18.43% to 25.60% | 8.16% | 7.45% to 8.93% |
| NPV | 99.54% | 98.65% to 99.84% | 99.75% | 98.30% to 99.96% |
| Accuracy | 86.26% | 83.65% to 88.60% | 55.07% | 51.50% to 58.60% |

AUC was evaluated using continuous COSA and OSTA, while the other parameters were evaluated using the optimal cutoff (COSA>0 and OSTA≤-4).

**Table 5.** The risk reclassification performance of COSA when compared to OSTA.

| Group | Category-less NRI |  |  | IDI |  |  |
| --- | --- | --- | --- | --- | --- | --- |
|  | Estimate | 95% CI | P-value | Estimate | 95% CI | P-value |
| Women aged<65 | 0.469 | 0.259-0.6791 | <0.001 | 0.028 | 0.013-0.044 | <0.001 |
| Women aged≥65 | 0.578 | 0.441-0.716 | <0.001 | 0.015 | 0.003-0.027 | 0.012 |
| Men | 0.73 | 0.4348-1.025 | <0.001 | 0.068 | 0.009-0.127 | 0.023 |

**Figure 1.**
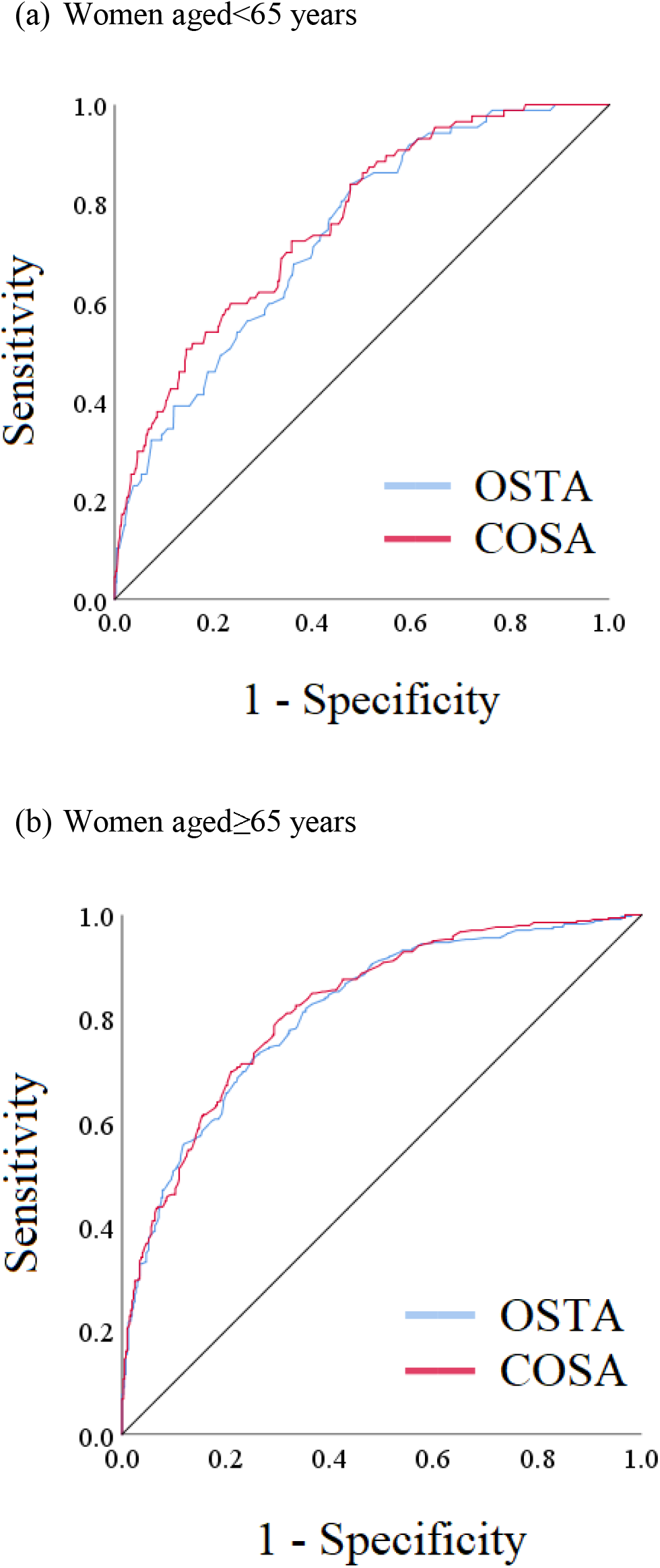
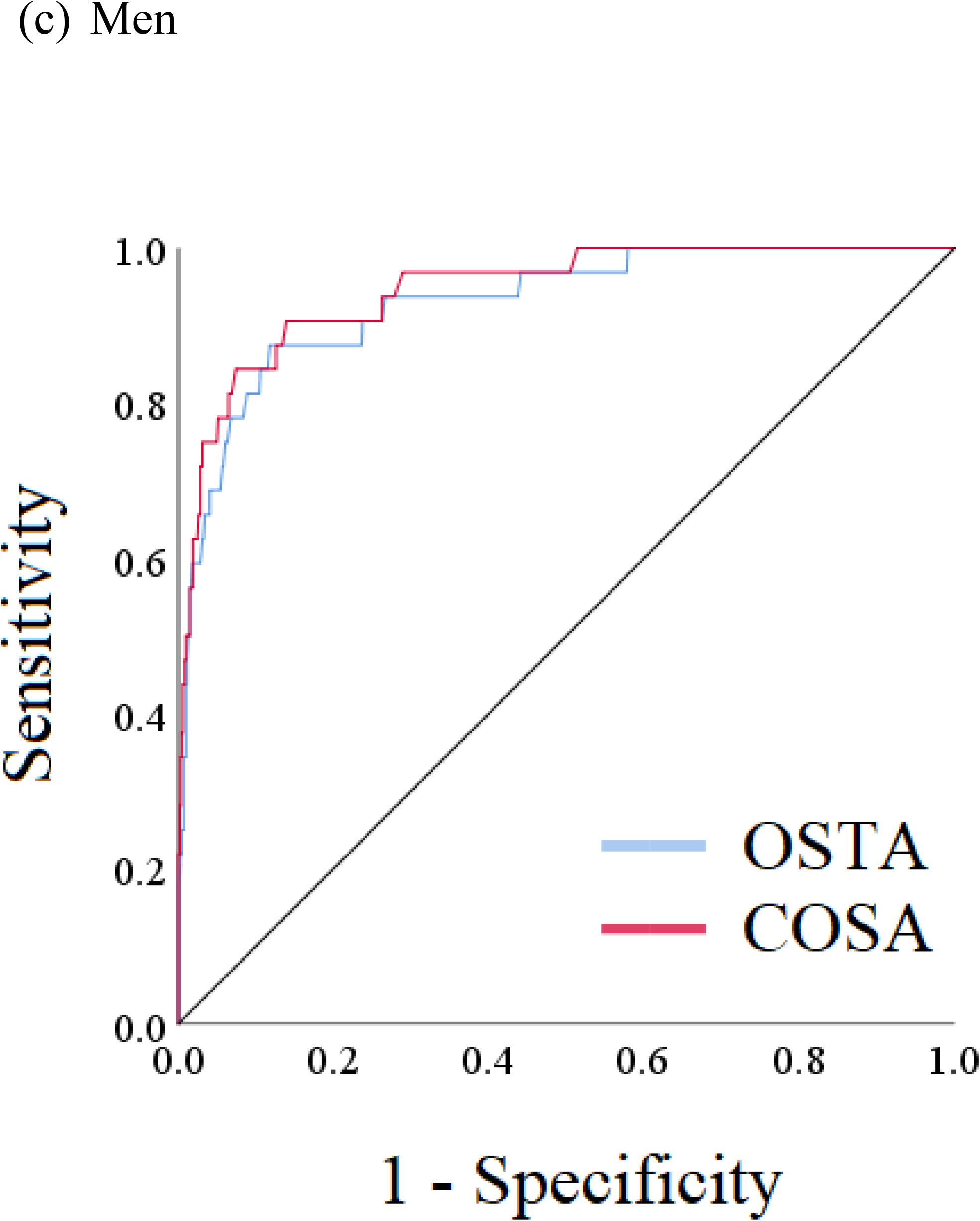
The AUC of COSA and OSTA in the internal validation cohort in (a) women aged<65 years, (b) women aged≥65 years, and (c) men.

### External validation of COSA

The PPV of the COSA was assessed in the external validation cohort comprising community-dwelling subjects. We screened 1,876 community dwelling participants (1,484 women and 392 men) without missing data (Supplementary Table 2). Among the 1,876 participants, 359 women and 60 men had the COSA>0 and were invited to have a DXA scan. A total of 116 women (32.3%) and 29 men (48.3%) declined to have a DXA scan, leaving 243 women and 31 men in the final analyses.

The DXA result of the subjects with COSA>0 is shown in Table 6. Among the 133 women aged <65, 54 (40.6%), 70 (52.6%), and 9 (6.8%) had osteoporosis, osteopenia, and normal BMD, respectively, resulting in a PPV of 40.6%. Among the 143 women aged≥65, 85 (59.4%), 51 (35.7%) and 7 (4.9%) had osteoporosis, osteopenia, and normal BMD, respectively, resulting a PPV of 59.4%. Among the 31 men, 6 (19.4%), 17 (54.8%), and 8 (25.8%) had osteoporosis, osteopenia, and normal BMD, respectively, resulting a PPV of 19.4%.

**Table 6.** Osteoporosis status in 274 community-dwelling subjects with COSA>0 and DXA scanned.

| Group | Osteoporosis |  | Osteopenia |  | Normal |  |
| --- | --- | --- | --- | --- | --- | --- |
|  | N | % | N | % | N | % |
| Women aged<65 | 54 | 40.6 | 70 | 52.6 | 9 | 6.8 |
| Women aged≥65 | 85 | 59.4 | 51 | 35.7 | 7 | 4.9 |
| Men | 6 | 15.8 | 20 | 52.6 | 12 | 31.6 |

## Discussion

In this study, we developed and validated the osteoporosis screening algorithm, COSA, in the Hong Kong Chinese population. We compared the performance of COSA with OSTA, and found that COSA had a significantly improved reclassification of osteoporosis when compared to OSTA. In an external validation cohort comprising1,876 community dwelling subjects recruited from 2019 to 2021, we showed that the PPV of COSA was 40.6% in women aged<65, 59.4% in women aged≥65, and 19.4% in men.

To our knowledge, there was only one widely validated osteoporosis screening algorithm in Asia, OSTA, which has been validated in Chinese, Indian, Singaporean, and Thai. Among postmenopausal women in Beijing, OSTA has a sensitivity of 69.9%, specificity of 75.1%, PPV of 52.5%, and NPV of 86.2%.^9^ For postmenopausal women in rural area of India, OSTA has a sensitivity of 88.5% and specificity of 41.7% in identifying osteoporosis at the femoral neck.^10^ In Singaporean, a modified cutoff point was used (−1.2) with sensitivity of 76%, specificity of 74%, PPV of 48%, and NPV of 91%.^11^ Among middle-aged pre- and early post-menopausal women in Thai, OSTA has a sensitivity of 57.3%, specificity of 76.8%, PPV of 16.34%, and NPV of 95.83%.^12^ The overall performance of OSTA in identifying subjects at risk of osteoporosis varied between population, which could be due to study design, rural versus non-rural subjects, age of subjects included, ethnicity, etc. In general, we observed the lowest specificity and highest sensitivity in women aged≥65 years when compared to other studies, which reflects that the cutoff point of −1 identify almost all osteoporosis cases but also include a large number of subjects without osteoporosis. Conversely, COSA has a significantly improved reclassification of osteoporosis in all groups, suggesting that COSA can be used to screen for osteoporosis in Hong Kong Chinese more accurately.

Population-specific risk factors are more important in predicting a clinical outcome. We previously showed that the risk score composed of Hong Kong specific risk factors outperformed the FRAX in predicting hip fracture.^13^ In the current study, we included a number of well-established risk factors in identifying subjects with osteoporosis and selected the most robust predictors that passed Bonferroni correction. These factors were consistently and independently associated with osteoporosis in both development and validation cohorts. These could potentially explain why COSA had a higher accuracy when compared to the OSTA, as OSTA was originally developed based on a group of Asian women with different ethnicities, the beta estimates of OSTA are more specific for the Asian women, which may be less generalizable to Chinese women. In addition, we incorporated into COSA the history of fracture, a strong risk factor of osteoporosis and fracture, which could further explain why COSA performed better than OSTA in identification of people with osteoporosis.

The real-world external validation study further demonstrated the usefulness of COSA in population screening of osteoporosis. The PPV of COSA in the internal and external validation was 21.8% vs 19.4% for men, 23.15% vs. 42.6% in women aged <65 years, and 73.46% vs 62% in women aged≥65 years. The PPV observed in women aged≥65 years was ∼10% lower than that observed in the internal validation cohort. One of the reasons could be explained by the overlap of recruitment period with the COVID-19 pandemic period, during which the Government advocated the elderly to stay home. The potential healthy cohort bias is supported by the lower percentage of participants with COSA>0 in women aged≥65 years, when compared to women aged<65 years (Supplementary Table 2). Therefore, we expect that the actual PPV of COSA in women aged≥65 years should be higher than that observed in the current study. In addition, we previously reported a secular increase in BMD in Hong Kong Chinese population.^14^ Since the HKOS cohort study was established from 1995 to 2010, it is expected that the BMD in Hong Kong population has been further improved, leading to a lower PPV.

The current study has important clinical implications. We recently reported that the global hip fracture burden is increasing even though declining trends in hip fracture incidence were observed in many countries. To reduce the absolute number of hip fracture in the future, more efforts should be put in reducing risk of hip fracture. One possible way is to implement population screening. A previous randomized controlled trial in the United Kingdom showed that a community-based screening intervention reduced hip fracture risk.^15^ Thus, our simple tool, COSA, can be used to screen for osteoporosis in the community. Given that DXA availability is limited, our tool can be used to prioritize high-risk subjects for DXA diagnosis. This can reduce the number of people needed for having DXA scan. Moreover, this simple tool can also improve public awareness of osteoporosis and easily implemented by the non-government organizations and used by the users themselves.

Our study has several strengths. The HKOS cohort is a well-established cohort study of osteoporosis with a large sample size. Thus, our finding should have a high generalizability to the Hong Kong population. We included both internal and external validation cohorts, allowing us to evaluate the usefulness of the tool in screening osteoporosis. Nevertheless, there are limitations. First, HKOS was established more than a decade ago as aforementioned, the average BMD in Hong Kong population should have been improved. Thus, the actual sensitivity and specificity should have changed. Further validation study is required to evaluate the actual accuracy of COSA using database or cohorts with more recently collected data. Second, COSA was developed for identifying people with high risk of osteoporosis, whether it can be used to predict fracture requires further study. Third, the generalizability to other population is unknown.

In conclusion, we developed a simple osteoporosis screening algorithm, COSA, for Hong Kong Chinese population. COSA had a significantly higher accuracy than the existing osteoporosis screening tool, OSTA. Using COSA in prioritizing subjects for DXA scan could prevent subsequent hip fracture and improve public awareness of osteoporosis.

## Data Availability

All data produced in the present study are available upon reasonable request to the authors.

## Contributors

CLC contributed to conceptualisation and study design, and act as guarantors for the study. CLC performed statistical analysis, implemented development and validation of prediction models, and drafted the manuscript. CLC, KCT and AWK contributed to the data resources. CM contributed to subject recruitment in the external validation. CLC, GHL, CM, KCT, and AWK provided critical input to the analyses and discussion. All authors contributed to the data interpretation, critically reviewed and revised the manuscript, and approved the final manuscript.

## Declaration of Interests

CLC reports grants and personal fees from Amgen outside the submitted work. The other authors have nothing to declare.

## Funding

The study was funded by Amgen Inc.

## Supplementary Figures

**Supplementary Figure 1.**
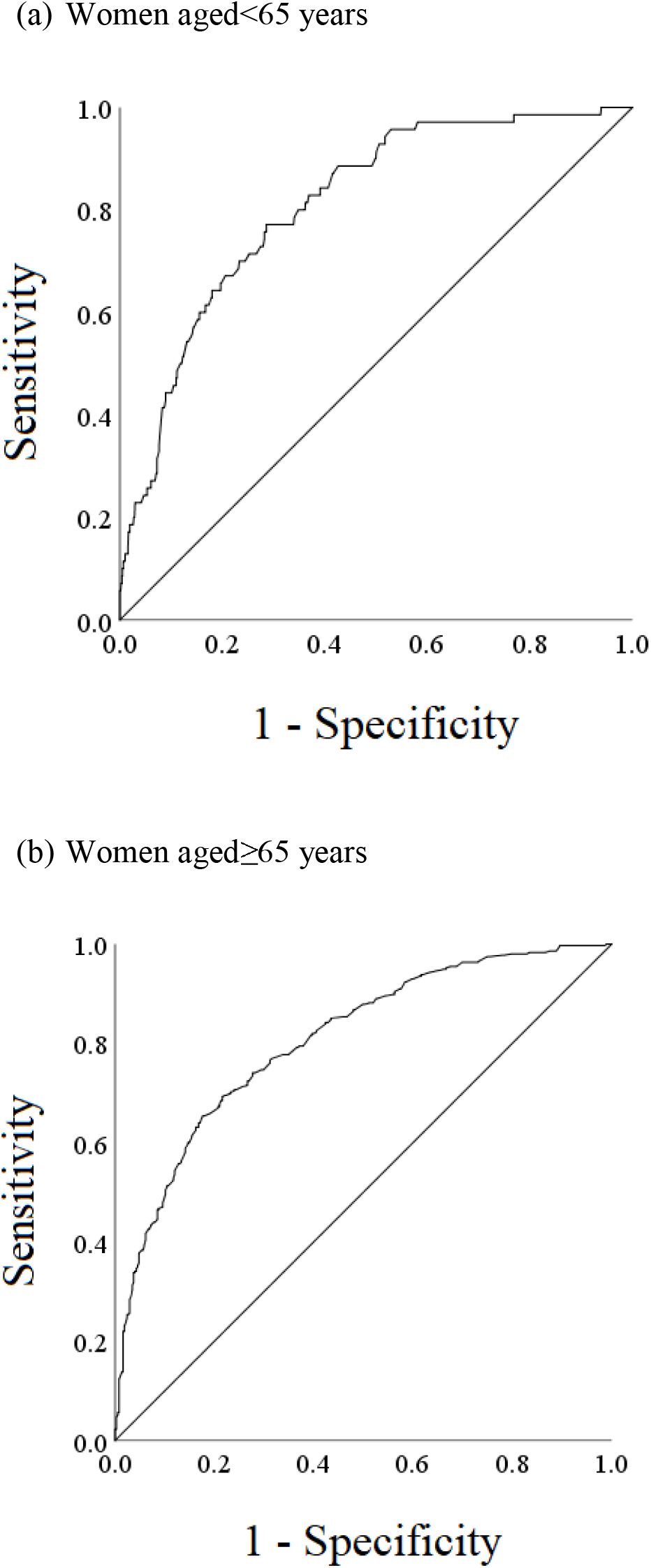
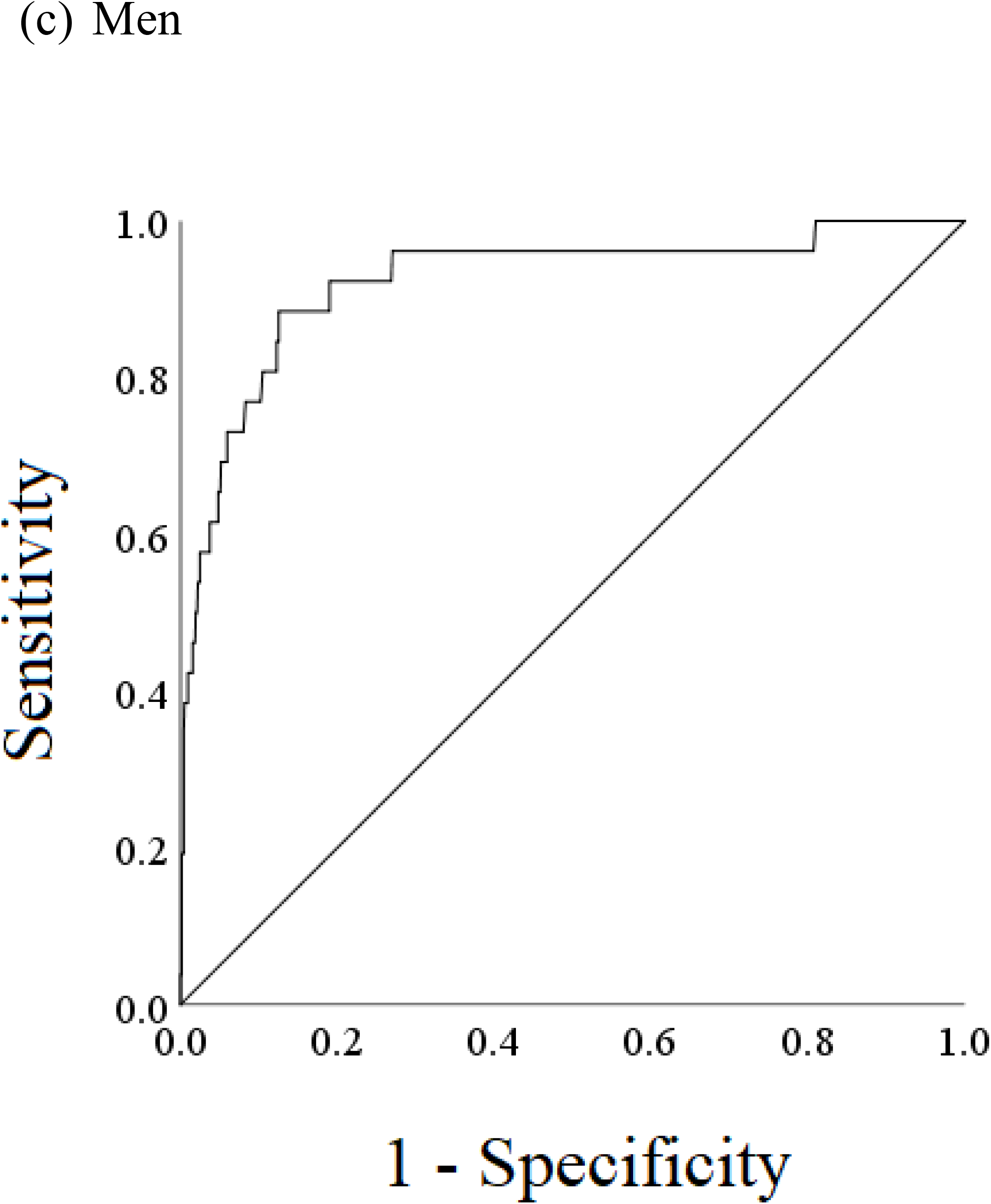
The AUC of COSA in the development cohort in (a) women aged<65 years, (b) women aged≥65 years, and (c) men.

**Supplementary Table 1.** AUC of the COSA in identification of osteoporosis at either spine or hip.

| Osteoporosis | Group | Development cohort |  |  |
| --- | --- | --- | --- | --- |
|  |  | AUC | 95% CI |  |
|  |  |  | lower | upper |
| Femoral neck | Women aged<65 years | 0.810 | 0.762 | 0.859 |
|  | Women aged>65 years | 0.807 | 0.776 | 0.838 |
|  | Men | 0.923 | 0.860 | 0.985 |
| Spine or hip | Women aged<65 years | 0.768 | 0.729 | 0.806 |
|  | Women aged>65 years | 0.790 | 0.757 | 0.823 |
|  | Men | 0.833 | 0.768 | 0.898 |
| All P<0.001 |  |  |  |  |

**Supplementary Table 2.** Demographic characteristics of the community-dwelling subjects.

|  | Women aged<65<br>(N=598) |  | Women aged≥65<br>(N=886) |  | Men (N=392) |  |
| --- | --- | --- | --- | --- | --- | --- |
|  | Mean<br>or N | SD<br>or % | Mean<br>or N | SD<br>or % | Mean<br>or N | SD<br>or % |
| Age (years) | 58.75 | 3.95 | 72.14 | 5.95 | 69.15 | 8.8 |
| Weight (kg) | 57.22 | 12.39 | 56.31 | 11.78 | 65.5 | 12.84 |
| History of fracture | 43 | 7.2 | 112 | 12.6 | 39 | 9.9 |
| COSA>0 | 151 | 25.3 | 208 | 23.5 | 60 | 15.3 |

Continuous variables are presented as mean and standard deviation (SD). Categorical variables are presented as number (N) and %.

